# The Neutrophil Lymphocyte Ratio (NLR), Platelet Lymphocyte Ratio (PLR) and routine hematological parameters of COVID-19 Patient: A perspective of the Indian scenario from a frontline pilot study of 32 COVID-19 cases in a Tertiary Care Institute of North India

**DOI:** 10.1101/2020.05.29.20102913

**Authors:** Neema Tiwari, Devajit Nath, Jyotsna Madan, Savitri Singh, Prashant Bajpai, Ujjwal Madan

**Author notes:** **Place of Study**: Department of Pathology, Super Speciality Pediatric Hospital & Postgraduate Teaching Institute, Sector 30, NOIDA 201301, India. **Address for correspondence:** Dr. Devajit Nath, Assistant Prof. Dept. of Pathology, Super Speciality Pediatric Hospital & Postgraduate Teaching Institute, Sector 30, NOIDA^1^ PIN-201301, India. Email: Dr.Neema Tiwari; Dr.Jyotsna Madan; Dr.Savitri Singh; Prashant Bajpai; Dr.Ujjwal Madan.

## Abstract

**Introduction:** The corona virus disease 2019(COVID-19) is caused by the virus SARS-CoV-2 and is declared as a global pandemic by World Health Organization (WHO). Various hematological parameters alteration has been documented in the Chinese literature in SARS-Cov-2 infection. However, there is a need for research to evaluate the pattern of the hematological parameters of COVID-19 patients in the Indian population.

**Aims & Objectives:** The objective of the study is to see the Neutrophil-Lymphocyte Ratio (NLR), Platelet Lymphocyte Ratio (PLR), and other hematological parameters alteration of COVID-19 patients along with their clinical course in the Indian scenario.

**Methods:** A single-center prospective study of 32 patients with laboratory-confirmed COVID-19 admitted to Super Speciality Pediatric Hospital & Post Graduate Teaching Institute NOIDA, from March to April, were enrolled for the study. The demographic date, the clinical status of the patients during admission and follow up, baseline, and follow up hematological findings were recorded. Statistical analysis of the data was carried out, and relevant findings were presented.

**Results:** Demographic characterization shows a mean age of 37.7 years, male (41.9%),female (58.1%)with majority patients are mildly symptomatic to asymptomatic(93%). The CBC values and NLR, PLR at baseline between the male and the female patients, are not showing any statistically significant difference as the 95% C.I. A statistically significant increment in the lab parameters is observed in follow-up visits.

**Conclusion:** Majority of the patients are younger and have mild clinical presentation with female predominance. Pediatric cases have mild symptomology. Baseline CBC findings show mild neutrophilia, lymphopenia, eosinopenia and normal to mild thrombocytopenia. An increase in CBC parameters, NLR was noted in follow up cases. Anemia was not noted in baseline CBC and in follow up group. A onetime PLR is not indicative of disease progression.

## INTRODUCTION

China reported a cluster of pneumonia cases of unknown etiology originating from “Wuhan, Hubei Province, China” to the WHO Country office on the 31^st^ of December 2019. The Etiology of the illness was attributed to a novel Virus belonging to the Corona Virus (CoV) Family. Subsequently, the World Health Organization officially announced that the disease caused by this new CoV is named COVID-19 (Corona Virus Disease-19).

Initially, the new Virus was called as 2019 nCovid. Subsequently, the Task of experts of the International Committee on Taxonomy of Viruses (ICTV) termed it as the SARS-COV-2 Virus as it is very similar to the one that caused the SARS outbreak (SARS-COVs).

The Virus is spreading around the World, thus assuming the dramatic features of a pandemic Emergency. On the 11^th^ March 2020, WHO declared the COVID-19 as a pandemic. India confirmed its first case on 30^th^ January 2020 in Kerala. The national capital of India, Delhi, and the National Capital Region (NOIDA) reported the first case on 5^th^ March 2020, and as of now till 22^nd^ April 2020, India has 15,859 active cases as per data from Ministry of Health and Family Welfare, Government of India website.

The Clinical characterization of COVID-19 has been broadly defined by WHO*^1^* with most of the confirmed COVID-19 cases have mild to moderate clinical presentation. However, there can be rapid deterioration for severe patients who will undergo respiratory failure, septic shock, or multiple organ dysfunction, threatening life. Efficient laboratory indicators for the disease severity, therapeutic response, and disease outcome have not been fully investigated. Early identification of indicators distinguishing severe patients from mild to moderate ones can facilitate faster medical intervention, thereby lowering the mortality rate and optimizing the case of the extremely strained medical recourses in developing countries like India. The guidelines of the National Health Commission of China for COVID-19 5^th^ Edition^2^ and the WHO interim Guidance^1^ currently recommended two laboratory parameters-normal/decreased numbers of leucocytes or decreased number of lymphocytes as one of the criteria for the diagnosis of COVID-19 infection. Various hematological parameters alteration has been documented in the current literature review in SARS-Cov-2 infection in the Chinese and the Western population, which signifies the severity and the prognosis of the patient outcome. However, there is a need for research to evaluate the pattern of the hematological parameters of COVID-19 patients in the Indian population.

### AIM

The primary objective of the study is to see the Neutrophil-Lymphocyte Ratio, Platelet Lymphocyte Ratio, and other hematological parameters alteration of COVID 19 Patients in the Indian scenario. The secondary objective of the study is to assess the follow up hematological parameters of the patients in order to obtain key indicators of disease progression and outcome that will provide guidance for subsequent clinical practice.

## METHOD

### Patient selection

Single Center prospective study of 32 patients with laboratory-confirmed COVID-19 admitted to Super Speciality Pediatric Hospital & Post Graduate Teaching Institute, from March to April, were enrolled for the study. The diagnosis of COVID-19 was according to the Ministry of Health and Family Welfare (MOHFW) Government of India (GOI) guidelines and confirmed by RTPCR performed on respiratory samples of the patient. All pediatric and adult cases were included in the study. Patients with chronic lung diseases, hematological disorders, liver disease, and malignancy on treatment were excluded from the study. Patients were divided into asymptomatic (mild) patient, and symptomatic(moderate) patients based on the Ministry of Health and Family Welfare (MOHFW) Government of India (GOI) Revised Guidelines on clinical management of COVID-19.

### Baseline data Collection

The demographic date, the clinical status of the patients during admission, and follow up and hematological findings were recorded. The samples for complete blood count (CBC) and peripheral smears for microscopy examination were collected on the day of admission. All patients didn’t receive any treatment before blood sampling. The samples were tested for complete blood count on 5-part hematology analyzer Abbott Cell Dyn Ruby, and values were noted. Peripheral blood smears were stained by Romanowsky stains, and findings were noted.

### Follow up

The patients whose clinical status remained unchanged were reexamined for laboratory indexes. The composite endpoint of the study was a clinical improvement, stability, or deterioration.

### Statistical Analysis

Dates are presented on means and standard deviation (SD). Differences in values between tested confidence intervals were calculated. Correlation analysis used a simple linear correlation. The optimal cutoff. Value, sensitivity, and specificity of NLR, PLR, and other CBC variables were determined, paired T-test was applied, and follow up data presented separately. All statistical analyses were carried out by SPSS 15.0 (SPSS Inc., Chicago, USA).

## RESULTS

### Demographic characterization

A total of 32 patients were admitted with a mean age of 37.7 years (4 months to 81 years), including males & Females. There were 29 cases in the mild group and 3 cases in the moderate/severe group. Patients in the moderate group were older than the cases in the mild category, with a case of an 81-year-old patient being recorded as the oldest in our institute. Females constitute 58.10%, and males constitute 41.9% of the study population. The median follow up time was of 2 weeks. All the paediatric cases under study were stable and asymptomatic, with only supportive treatment. Adult patients included in the study were given supportive treatment, and only one case showed clinical deterioration over a period of 14 days of admission.

The statistical analyses of the data are being summarized in the table and graphs.

**Table 1:**
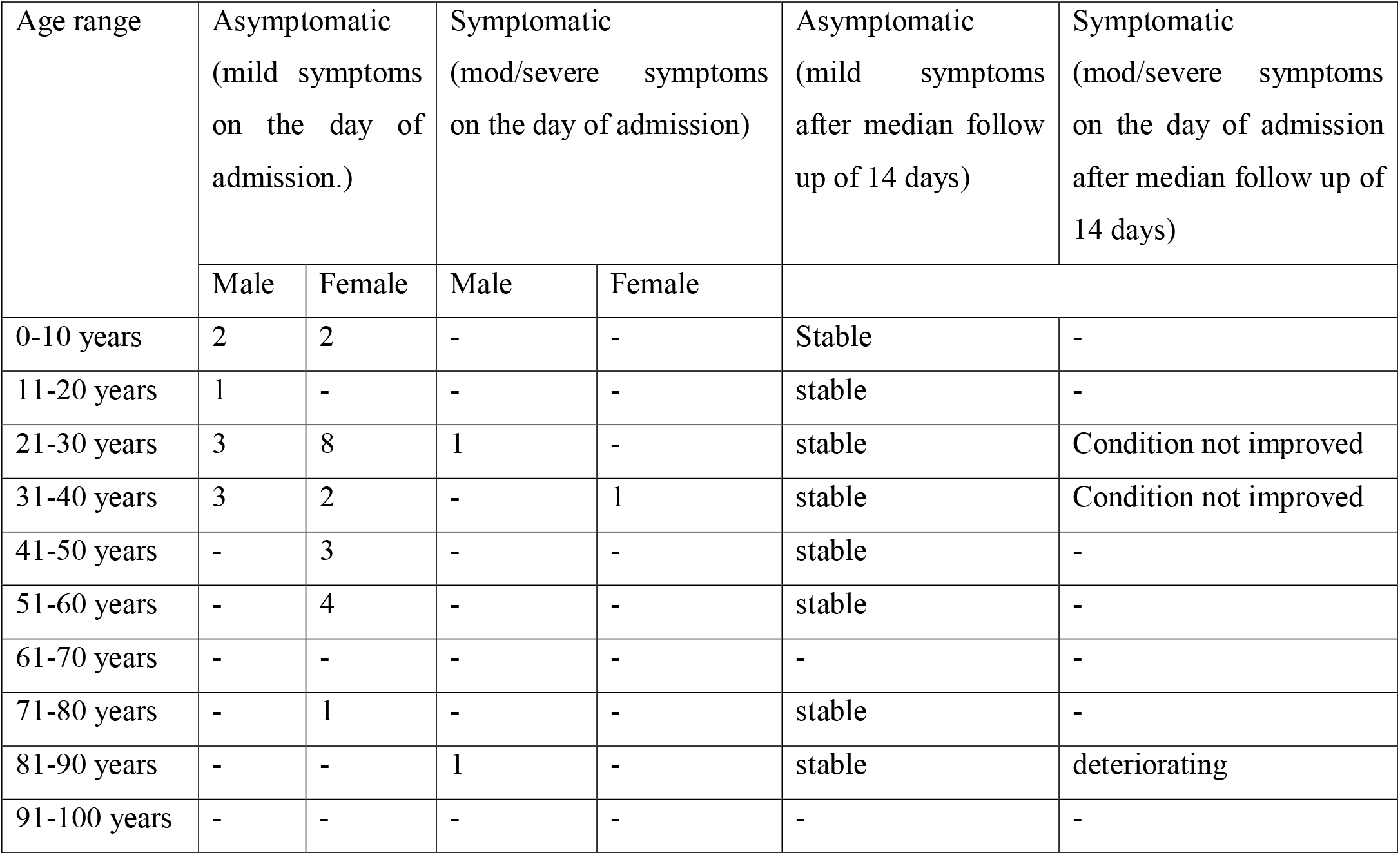
Demographic and Clinical data on the day of admission and after a median follow up of 14 days.

| Age range | Asymptomatic<br>(mild symptoms<br>on the day of<br>admission.) |  | Symptomatic<br>(mod/severe symptoms<br>on the day of admission) |  | Asymptomatic<br>(mild symptoms<br>after median follow<br>up of 14 days) | Symptomatic<br>(mod/severe symptoms<br>on the day of admission<br>after median follow up of<br>14 days) |
| --- | --- | --- | --- | --- | --- | --- |
|  | Male | Female | Male | Female |  |  |
| 0-10 years | 2 | 2 | - | - | Stable | - |
| 11-20 years | 1 | - | - | - | stable | - |
| 21-30 years | 3 | 8 | 1 | - | stable | Condition not improved |
| 31-40 years | 3 | 2 | - | 1 | stable | Condition not improved |
| 41-50 years | - | 3 | - | - | stable | - |
| 51-60 years | - | 4 | - | - | stable | - |
| 61-70 years | - | - | - | - | - | - |
| 71-80 years | - | 1 | - | - | stable | - |
| 81-90 years | - | - | 1 | - | stable | deteriorating |
| 91-100 years | - | - | - | - | - | - |

**Table 2:** Statistical analysis of haematological parameters of 32 cases and their correlation with each other for one time CBC collected on the day of admission.

| Female | Mean | median | Std deviation | 95% CI | Paired t test |
| --- | --- | --- | --- | --- | --- |
| RBC(3.6-4.69)$\times 10^6$ μL | 4.68 | 4.67 | 0.60 | Not significant | No significant correlation |
| Hb(10.8-14.2)g/dL | 12.45 | 12.35 | 2.06 | Not significant | No significant correlation |
| TLC(3.70-10.1)$\times 10^3$ μL | 6.81 | 6.34 | 1.71 | Not significant | No significant correlation |
| Absolute Neutrophil Count (1.63-6.96) $\times 10^9$ /L | 3.79 | 3.42 | 1.33 | Not significant | No significant correlation |
| Absolute Lymphocyte Count(1.09-2.99) $\times 10^9$ /L | 2.43 | 2.47 | 0.81 | Not significant | No significant correlation |
| Absolute Monocyte count(0.240-0.790) $\times 10^9$ /L | 0.41 | 0.32 | 0.21 | Not significant | No significant correlation |
| Absolute Eosinophil Count(0.030-0.440) $\times 10^9$ /L | 0.13 | 0.09 | 0.14 | Not significant | No significant correlation |
| Platelet count(155-366) $\times 10^3$ μL | 155 | 171 | 73 | Not significant | No significant correlation |
| NLR | 1.85 | 1.45 | 1.43 | Not significant | No significant correlation |
| PLR | 50.27 | 60.60 | 41.39 | Not significant | No significant correlation |
| ESR mm AEFH | 33 | 36 | 12 | Not significant | No significant correlation |
| Male | Mean | median | Std deviation | 95% CI | Paired t test |
| RBC(3.6-4.69)$\times 10^6$ μL | 5.10 | 5.19 | 0.74 | Not significant | No significant correlation |
| Hb(10.8-14.2)g/dL | 12.78 | 12.90 | 2.16 | Not significant | No significant correlation |
| TLC(3.70-10.1)$\times 10^3$ μL | 8.49 | 7.70 | 2.94 | Not significant | No significant correlation |
| Absolute Neutrophil Count (1.63-6.96)$\times 10^9$ /L | 4.92 | 4.37 | 2.55 | Not significant | No significant correlation |
| Absolute Lymphocyte Count(1.09-2.99) $\times 10^9$ /L | 2.9 | 2.48 | 2.05 | Not significant | No significant correlation |
| Absolute Monocyte count(0.240-0.790) $\times 10^9$ /L | 0.41 | 0.39 | 0.24 | Not significant | No significant correlation |
| Absolute Eosinophil Count(0.030-0.440) $\times 10^9$ /L | 0.09 | 0.09 | 0.07 | Not significant | No significant correlation |
| Platelet count(155-366) $\times 10^3$ μL | 197 | 163 | 103 | Not significant | No significant correlation |
| NLR | 2.13 | 1.85 | 1.47 | Not significant | No significant correlation |
| PLR | 39.65 | 40.90 | 33.40 | Not significant | No significant correlation |
| ESR mm AEFH | 33 | 35 | 5 | Not significant | No significant correlation |

The lab values at baseline are not showing any statistically significant difference as the 95% Confidence Interval (C.I). There is no statistically significant difference in the lab parameters between males and females at the baseline visit. The baseline Lab values of Follow-up cases are also not showing any statistically significant difference as the 95% Confidence Interval (C.I) for the mean of lab parameters are falling within the acceptable lab ranges (Normal ranges as per 5 part Hematology Analyzer Reference Value within the parentheses)

**Table 3:** Statistical analysis of the follow-up CBC data available for 3 cases in the symptomatic (moderate to severe symptoms) group.

| **3 cases** | **Mean** | **median** | **Std deviation** | **95% CI** | **Paired t test** |
| --- | --- | --- | --- | --- | --- |
| Age | 48 | 33 | 29 | - | - |
| RBC(3.6-4.69)$\times 10^6$ μL | 5.04 | 5.0 | 0.40 | Not Significant | Not significant |
| Hb(10.8-14.2)g/dL | 13.2 | 12.8 | 1.5 | Not Significant | Not significant |
| TLC(3.70-10.1)$\times 10^3$ μL | 10.4 | 10.5 | 3.4 | Significant increase | significant |
| Absolute Neutrophil Count (1.63-6.96) $\times 10^9$ /L | 5.9 | 4.3 | 4.05 | Significant increase | significant |
| Absolute Lymphocyte Count(1.09-2.99) $\times 10^9$ /L | 3.7 | 4.3 | 1.73 | Significant decrease | significant |
| Absolute Monocyte count(0.240-0.790) $\times 10^9$ /L | 0.6 | 0.5 | 0.34 | Significant increase | significant |
| Absolute Eosinophil Count(0.030-0.440) $\times 10^9$ /L | 0.05 | 0.017 | 0.08 | Significant decrease | significant |
| Platelet count<br>Platelet count(155-366) $\times 10^3$ μL | 283 | 154 | 227 | Significant increase | significant |
| NLR | 3.48 | 3.13 | 2.48 | Significant increase | significant |
| PLR | 118.8 | 40.9 | 171.11 | Significant | significant |
|  |  |  |  | increase |  |
| ESR<br>mmAEFH | 34 | 32 | 5 | Significant<br>increase | significant |

A statistically significant increment in the lab parameters is observed in follow-up visit 2 & visit 3, but a larger follow-up data needs to be studied for better delineation of the results (N=3 (Follow-up visit 2) & N=2 (Follow-up Visit 3)).

**Table 4:** The NLR in our study group asymptomatic/mild versus symptomatic/moderate-severe symptoms was as given in the table:

| Age range | NLR in 29 Asymptomatic (mild symptoms on the day of admission.) |  | NLR in 3 Symptomatic (mod/severe symptoms on the day of admission) |  | NLR in the 3 Symptomatic (mod/severe symptoms on 14<sup>th</sup> day follow up) |  |
| --- | --- | --- | --- | --- | --- | --- |
|  | <3.1 | >3.1 | <3.1 | >3.1 | <3.1 | >3.1 |
| <0-10 years | 3 | - | - | - | - | - |
| 11-20 years | - | - | - | - | - | - |
| 21-30 years | 7 | 1 | - | - | - | - |
| 31-40 years | 9 | 2 | - | - | - | - |
| 41-50 years | 2 | - | - | - | - | - |
| 51-60 years | 1 | - | - | 2 | - | 2 |
| 61-70 years | 2 | - | - | - | - | - |
| 71-80 years | 2 | - | - | - | - | - |
| 81-90 years | - | - | - | 1 | - | 1 |
| 91-100 years | - | - | - | - | - | - |

In our analysis, 6 cases were seen with baseline NLR above the standard risk stratification cut off value of 3.13. This was not statistically significant. However, out of the 6 cases, 3 cases were symptomatic with high NLR both in baseline CBC and on follow up after 14 days, which was statistically significant.

**Image 1 & Image 2:**
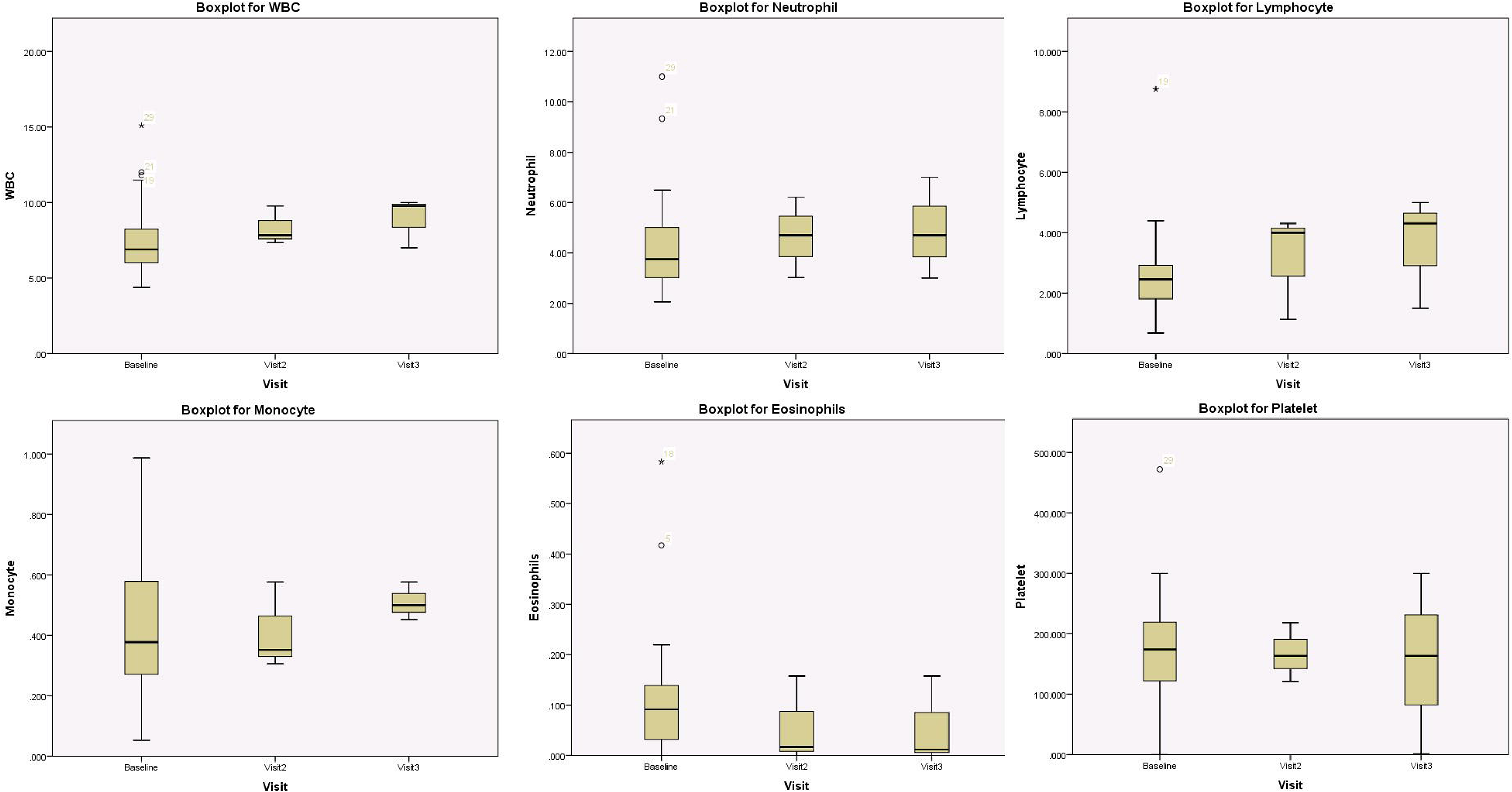
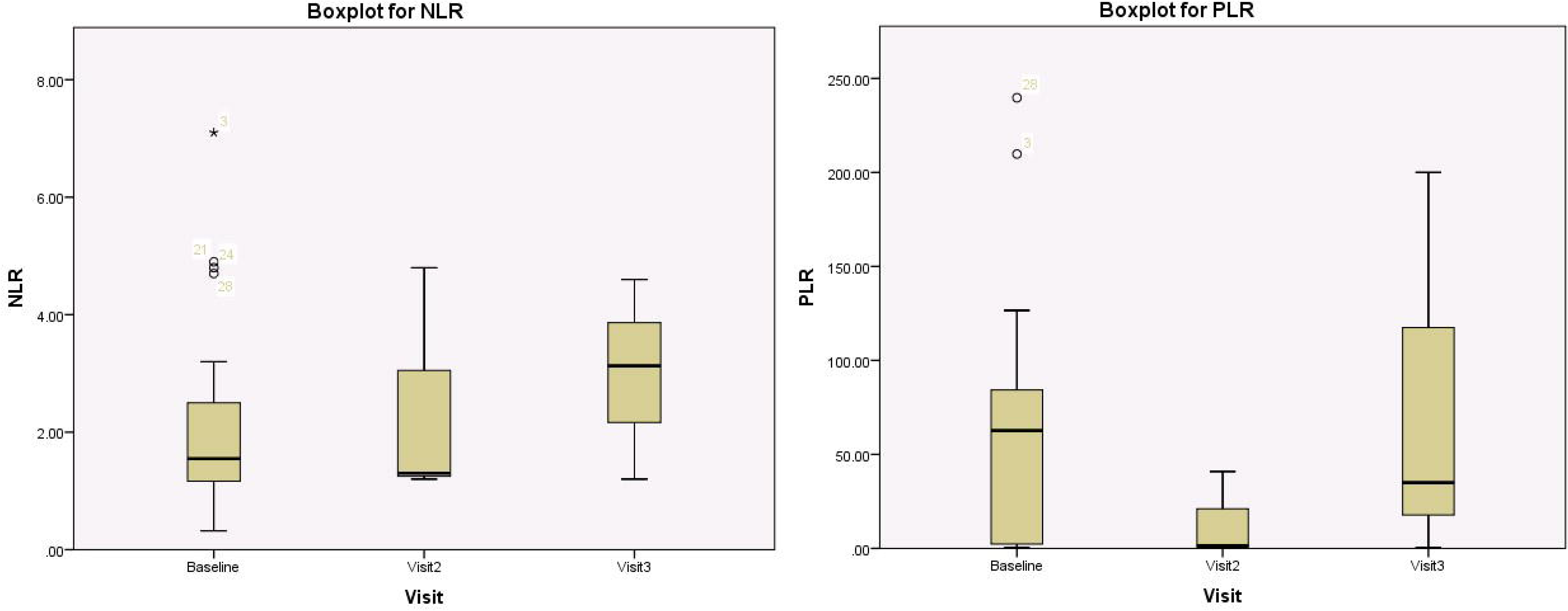
Composite analysis of variables under study for baseline values with follow up (box and whisker plots) show range of variation in different parameters. The lines show variation beyond the upper and lower ranges with each plot with random outliers. Each plot has a central line showing the range of dispersion of values and in the composite plots it is seen that compared to baseline there is a significant variation in the follow up plots for all the variables of CBC except Hemoglobin and RBC Count along with NLR and PLR.

**Image 3:**
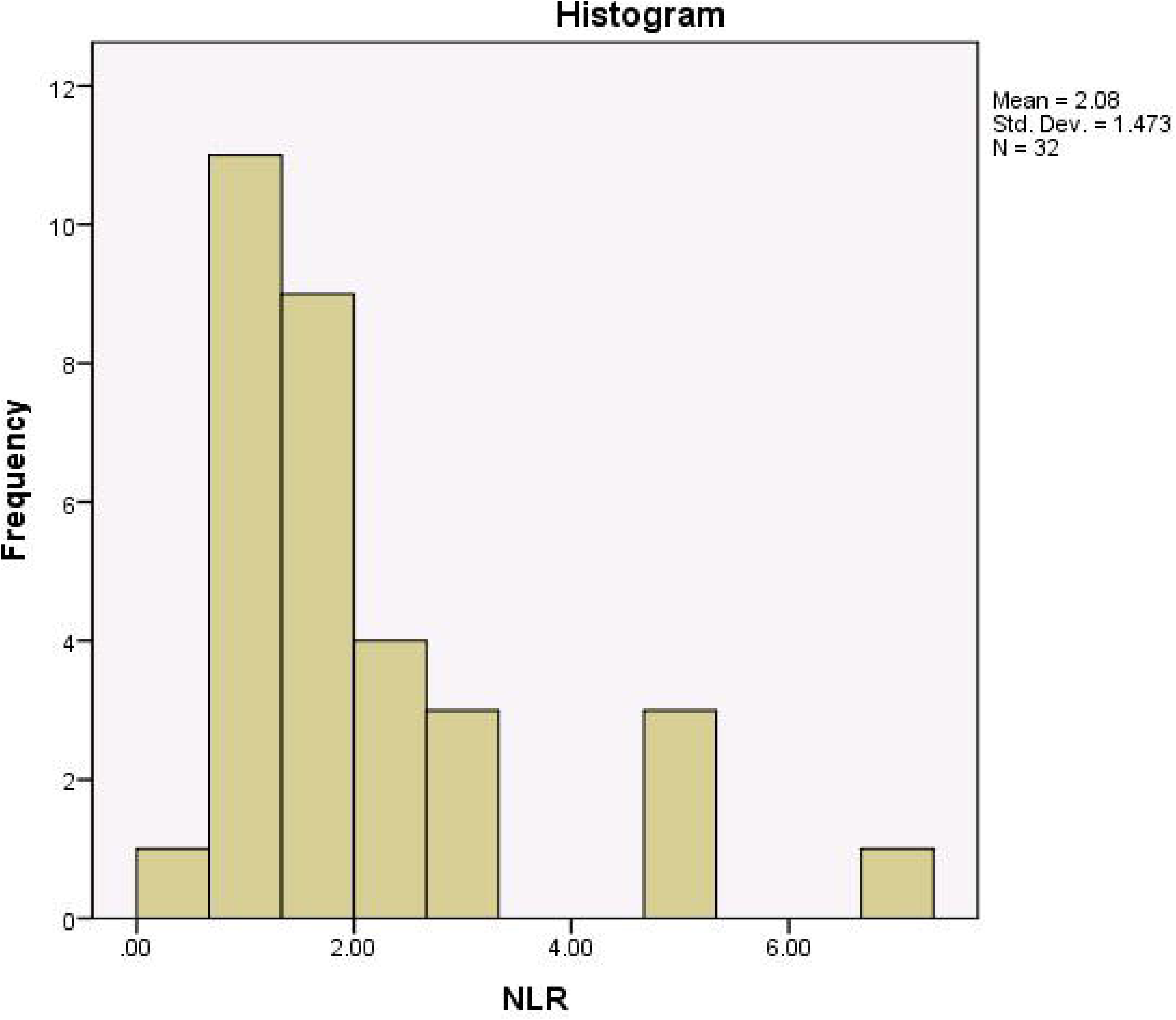
Baseline NLR histogram showing range variability of NLR in 32 cases with mean value of 2.08 with few values above 3.1.

**Image 4:**
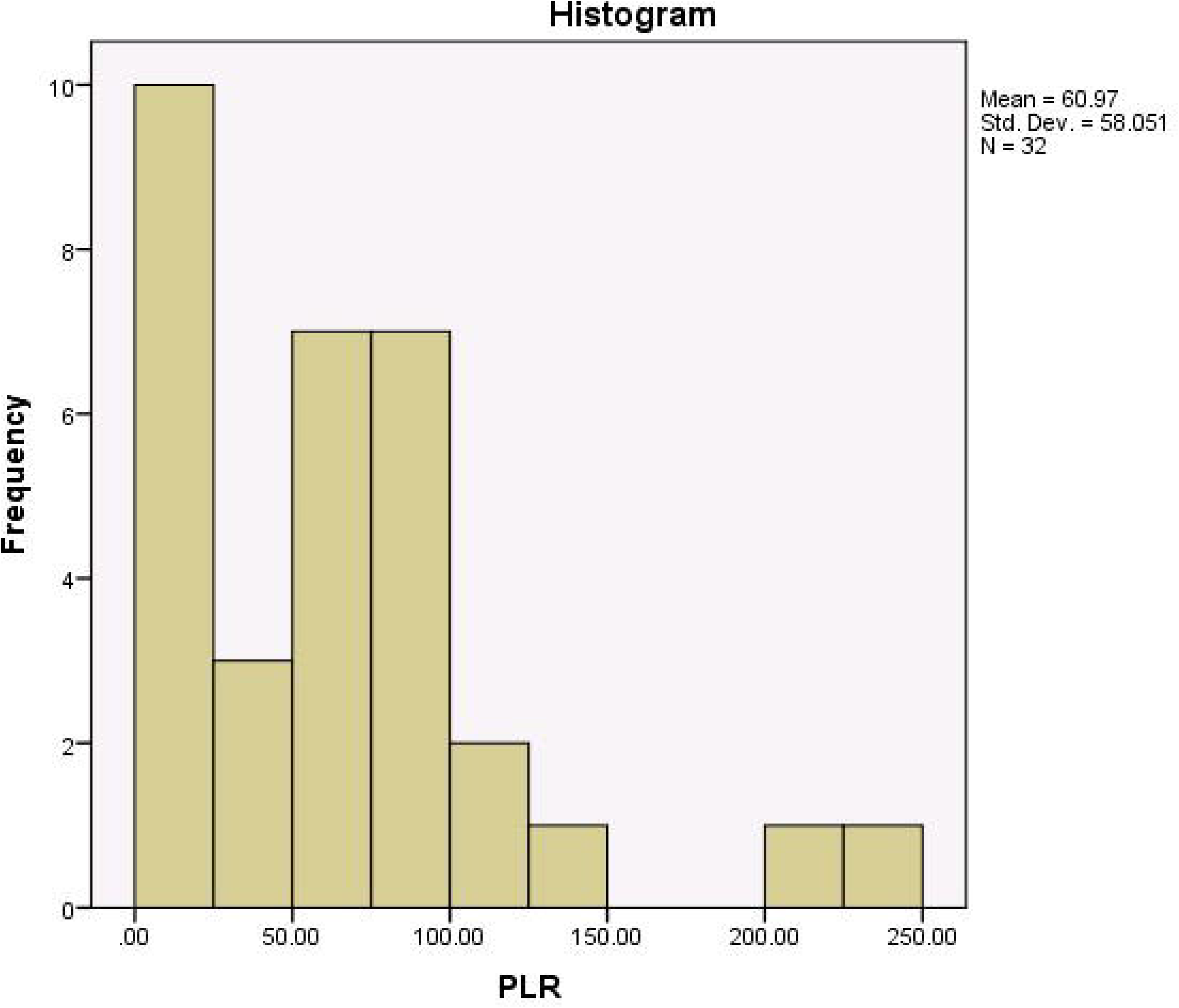
Histogram showing variability in the PLR range of 32 baseline cases with mean value of 60.97.

From the above statistical analysis, it can be presumed that the longer the virus remains in the human body, the CBC parameters are affected, but in the earlier phase of onset of disease or symptoms, significant alteration of the CBC parameters are not observed.

## DISCUSSION

Coronavirus disease 2019 (COVID-19) is a respiratory tract infection caused by a newly emergent coronavirus, SARS-CoV-2, that was first recognized in Wuhan, China, in December 2019. Genetic sequencing of the virus suggests that SARS-CoV-2 is a beta-coronavirus closely linked to the SARS virus^1^. While most people with COVID-19 develop a mild or uncomplicated illness, approximately 14% develop severe disease requiring hospitalization and oxygen support, and 5% require admission to an intensive care unit. Several studies have documented SARS-CoV-2 infection in patients who never develop symptoms (asymptomatic) and in patients not yet symptomatic (pre-symptomatic) *^3-7^*

Since asymptomatic persons are not routinely tested, the prevalence of asymptomatic infection and detection of pre-symptomatic infection is not well understood.

Clinical and epidemiological data of the asymptomatic and mildly symptomatic cases in Indian lacks to provide an illustration of the epidemiological curve of disease outbreak in the Indian population. Data from our study shows that most of the patients are asymptomatic to mildly symptomatic (90.3%), thus correlating with other studies in the Chinese and the Western population.

The demographic characteristics of our study showed that females were affected more often than men. This is in contrast to various studies conducted by Guan WJ et al*^8^*, Zhang JJ et al*^9^*, Nanshan Chen et al*^10^*, where the disease incidence was more in men as compared to females. Previous studies conducted on SERS COV and MERS have also been found to infect more males than females*^11,12^*. The reduced susceptibility of females to viral infections could be attributed to the protection from X chromosome and sex hormones, which play an important role in innate and adaptive immunity*^13^*.

The change in the gender pattern in the Indian population could be due to immunological, ethnic, or racial variation. This area needs further research, and a larger cohort study may be suggested to establish the fact.

A study conducted by Li et al. on the first 425 cases of COVID 19 in Wuhan; China showed that the patients’ median age was 59 years, with a range of 15 to 89 years*^14^*. However, in our study, the median age was 37.7 years (4 months-81 years). In our study group, pediatric cases comprise 12.9% of the total cases. This is in contrast to the study conducted by Li et al. in the Chinese population whereby they reported no clinical cases in children below 15 years of age*^14^*.

Although the clinical characteristic of COVID-19 has been broadly defined by WHO^1^, an outline of the most representative laboratory abnormalities found in patients with COVID-2019 infection still lacks in the Indian scenario.

Lippi G et al*^15^* carried out a systematic literature review and highlighted that the most important hematological parameter abnormalities observed in COVID-19 patients, which may predict the progression toward severe or critical forms of COVID-19, include leukocytosis, neutrophilia, and lymphopenia. Each of these prognostic parameters retains a specific clinical and biological significance, which, altogether, can contribute to reflect the evolution toward more unfavorable clinical pictures.

Huang C et al. and Yang X mentioned in their articles whereby 85% of the critically ill patients of their study group with COVID-19 showed lymphopenia *^16,17^*. The presence of lymphopenia as a signature of severe COVID-19 was confirmed by Bai et al., who reported that ICU patients suffering this infection had a median lymphocyte count of 800 cells/mm*^18^* with -non-survivors exhibiting persistent lymphopenia.

As per the IFCC guidelines, the CBC parameters in a COVID case show neutrophilia, leucocytosis lymphopenia, and thrombocytopenia (IFCC Information Guide on COVID-19; published Thursday, March 26: https://www.ifcc.org/ifcc-news/2020-03-26-ifcc-information-guide-on-covid-19/). In another Chinese study, which has analyzed data from two laboratory parameters, “normal/ decreased number of leukocytes” or “decreased number of lymphocytes,” as one of the criteria for the diagnosis of COVID-19 infection. A study conducted by Chuan Qin et al.*^19^* showed that primary dysregulation of the immune response, especially T lymphocytes, might be highly involved in the pathological process of COVID-19. Most of the severe cases demonstrated elevated levels of infection-related biomarkers and inflammatory cytokines. The number of T cells significantly decreased, and more hampered in severe cases*^18^*.

Analysis of the baseline CBC parameters of our study population showed that 4 cases (12.9%) showed neutrophilia, 3(9.6%) cases showed lymphopenia (ALC < 1 xJ10^9^/L), and 5 cases (16.1%) showed monocytosis. However, the baseline total leucocyte count was not increased. The lower percentage of cases showing lymphopenia in our study is in contrast to the other studies conducted in China, whereby 63% of cases showed lymphopenia and 42% cases outside the Chinese population*^15,19^*. The disparity in the percentage of cases showing lymphopenia may, in part, be reflective of the epidemiological variation of the Indian population. The finding of monocytosis in our study has not been documented in other studies.

Out of the 32 cases, 8 cases (25.8%) had eosinopenia. Our findings matched with a single Chinese study, which also highlights eosinopenia as a significant prognosticating factor*^20^*. The study proposes Eosinopenia as a potentially more reliable laboratory predictor of SARS-CoV-2 infection than recommended “leukocyte counts” and “lymphopenia”*^20^*

From a study conducted in Huizhou, China of the 30 COVID 19 cases, it was concluded that patients with markedly elevated platelets and longer average hospitalization days might be related to the cytokine storm. The Platelet Lymphocyte Ratio (PLR) of the patients means the degree of cytokine storm, which might provide a new indicator in the monitoring in patients with COVID-19*^21,22^*. In our study, we had 8 cases (25.8 %) with thrombocytopenia, and only one case (3.2%) had mild thrombocytosis. However, the relevance of platelet count and PLR can only be interpreted if follow up samples at different time points in a large cohort are available.

The Neutrophil-to-Lymphocyte ratio (NLR) is a biomarker for the assessment of the severity of bacterial infections and the prognosis of patients with pneumonia and tumor.NLR may serve as a surrogate marker of early risk identification in COVID 19 infection. In a study by Forget et al.*^23^*, it was identified that normal NLR values in an adult, non-geriatric population in good health are between 0.78 and 3.53. Whereas, in another study by Jingyuan Liu*^24^*, it was concluded that Patients with age ≥50 having NRL ≥3.13 are at risk of severe illness, and they should get rapid access to the intensive care unit, if necessary.

In our analysis, 6 cases were seen with baseline NLR above the cut off 3.13. This was not statistically significant. However, out of the 6 cases, 3 cases were symptomatic with high NLR both in baseline CBC and on follow up after 14 days, which was statistically significant.

In an accepted article to the editor, Bingwen Eugene Fan et al*^25^* mentioned that in their study population in Singapore, microscopic peripheral blood film examination shows a higher number of patients who are lymphopenic & have the presence of a few reactive lymphocytes, a subset of which appears to be lymphoplasmacytoid. Peripheral blood smear examination in our cases showed a predominantly normocytic normochromic picture with few cases showing toxic granules in neutrophils while occasional case showed activated lymphoid cells, the finding which was matching with that of Bingwen Eugene Fan et al. study*^25^*.

This finding is in contrast with the study conducted during the Severe Acute Respiratory Syndrome (SARS) outbreak in 2003*^26^*, where reactive lymphocytes were not observed in the peripheral blood film examination of the SARS patients in Singapore and only in 15.2% of cases in a similar study in Hong Kong*^27^*.

From the study, it could be concluded that baseline CBC parameters of all the 32 cases presenting with mild to moderate symptoms did not show any statistically significant variation in the parameters that may substantially mark their clinical progress or severity. Also, there is no statistically significant difference in the lab parameters between males and females at the baseline visit. One reason for this observation could be due to the limited sample size or early /asymptomatic/mild presentation of COVID cases.

In the follow-up group, the lab values at Follow-up visit one are not showing any statistically significant difference in the CBC parameters. A statistically significant increment in the lab parameters like TLC, neutrophils, monocytes was seen while increasing lymphopenia, and developing eosinopenia was also seen in is follow-up visit 2 & visit 3. Platelet count and hemoglobin levels remained unchanged even with disease progression in our study. The statistically significant increment in the CBC parameters in this follow up group was correlating with the patient clinical status. A small follow up group is a statistical limitation of our study. A follow-up analysis of CBC Parameters of a large Indian cohort is suggested to statistically prove the association of parameters with prognosis.

In a recently discussed paper, it is suggested that in addition to various mechanisms, SARS-CoV-2 viral protein infects hemoglobin by the immune hemolysis of red blood cells*^28^*.

However, in our study, the baseline RBC count and hemoglobin levels were not changed in all the 32 cases and in the 3 follow up cases on 1^st^, 2^nd^, 3^rd^ reading.

These statistically insignificant findings of baseline CBC in Indian population with a stable clinical course as compared to other foreign data are promising and could explain worse COVID-19 outcome in western countries, unlike India which is leading globally in disease containment with only 12759 cased,1515 recoveries and only 420 deaths compared to world data of 1,40,773 deaths (0.29% of the world deaths). However, a relevant trend in follows up CBC in a large Indian cohort might predict the disease progression and prognosis.

### Limitation of the Study

There may be missing laboratory data as CBC investigation was not performed daily on all patients, especially those who are minimally symptomatic and improving in the general Isolation Ward. Also, a very low sample size of follow up as well as no follow up cases analyzed is a big limitation for definitive statistical findings.

## CONCLUSION

The study of 32 COVID-19 cases of Indian population shows that majority of the patients are younger, have asymptomatic to mild clinical presentation, and a higher incidence in the female population. The majority of pediatric cases have mild symptomology with a stable clinical course.

- Baseline CBC findings of all the cases show mild neutrophilia, mild lymphopenia, eosinopenia, mild monocytosis, and a normal to mild thrombocytopenia. However, the statistical distributions of the parameters were within the normal range without any association of disease progression. Peripheral blood smear examination in our cases showed a predominantly normocytic normochromic picture with few cases showing toxic granules in neutrophils while the occasional case showed activated lymphoid cells.
- A significant statistical trend of increase in CBC parameters, NLR, was noted in follow up cases with persistent symptoms; however, a larger follow-up cohort is needed to arrive at a statistical significance.
- Anemia was not noted in baseline CBC and in the follow-up group.
- A onetime PLR is not indicative of disease progression.

## Data Availability

THIS IS A PROSPECTIVE STUDY DONE ON DATA AVAILABLE FROM ROUTINE INVESTIGATIONS

## Acknowledgement

We thank the Department of Pediatric Medicine, SSPHPGTI, NOIDA, India and the Internal Medicine, Government District Hospital, NOIDA, India for their support in carrying out the study and the technical staff, Department of Pathology, SSPHPGTI, NOIDA for their effort in carrying out the laboratory investigations. Special acknowledgment to Jahnu Saikia, Ph.D. Scholar, Department of Biosciences and Bioengineering, Indian Institute of Technology Guwahati, India for his contribution in supervising the final proofreading and the references of the manuscript. We also greatly appreciate the efforts of healthcare workers and the support of their families during this outbreak

### Research in context

#### Evidence before this study

Current literature review documents the epidemiological and hematological parameter alteration of the COVID-19 patients in the Chinese and the western population, thereby perpetuating a knowledge deficit in the Indian population where the pandemic is spreading over time.

#### Added value of this study

The study obtained data from 32 COVID-19 cases in NOIDA, India to explore the epidemiology and the baseline and follow up hematological parameters of the patients. The pilot study provides further information on the demographic and laboratory hematological parameters of the patients thereby adding substantial inputs to the knowledge data base.

#### Implication of the outcome of the present study

The present study shows that in India majority of the cases have asymptomatic to mild clinical presentation with female outnumbered the male and a majority of pediatric cases have mild symptomology with stable clinical course. No statistically significant baseline hematology parameters alteration was noted in the study group however follow up cases show statistical significance in the parameters.

The slightly different epidemiological and hematological pattern of the Indian cases will help the clinicians and the researchers for better understanding of the disease pattern with timely management of the epidemic.

## Author Declarations

### Ethics committee approval

All relevant ethical guidelines have been followed; any necessary IRB and/or ethics committee approvals have been obtained and details of the IRB/oversight body are included in the manuscript-Yes.

The details of the IRB/oversight body that provided approval or exemption for the research described are given below:

Institutional Ethics Committee [IEC] Super Speciality Paediatric Hospital & Post Graduate Teaching Institute,NOIDA 201301,India. Standard Operating Procedures of Institutional Ethics Committee of SSPHPGTI July, 2019- Annexures: AM-V1/SOP-13/V1/AN2-V1/SOP-13/V1 Website: www.sspginoida.ac.in

All necessary patient/participant consent has been obtained and the appropriate institutional forms have been archived-Yes

Institutional Ethics Committee [IEC] Super Speciality Paediatric Hospital & Post Graduate Teaching Institute,NOIDA 201301,India. Standard Operating Procedures of Institutional Ethics Committee of SSPHPGTI July, 2019. Website: www.sspginoida.ac.in

I understand that all clinical trials and any other prospective interventional studies must be registered with an ICMJE-approved registry, such as ClinicalTrials.gov. I confirm that any such study reported in the manuscript has been registered and the trial registration ID is provided (note: if posting a prospective study registered retrospectively, please provide a statement in the trial ID field explaining why the study was not registered in advance)-Yes

I have followed all appropriate research reporting guidelines and uploaded the relevant EQUATOR Network research reporting checklist(s) and other pertinent material as supplementary files, if applicable-Yes

## Author contribution

Authors of the manuscript declare contribution in the study

## COVER LETTER

Respected Sir/Madam,

This is to request you to kindly consider my manuscript/original article titled’ **The Neutrophil Lymphocyte Ratio (NLR), Platelet Lymphocyte Ratio (PLR) and routine hematological parameters of COVID-19 Patient: A perspective of the Indian scenario from a frontline pilot study of 32 COVID-19 cases in a Tertiary Care Institute of North India.’** In your esteemed platform for publication. There is no conflict of interest and all authors have contributed in the formulation of this manuscript. This manuscript has only been communicated to this platform at present.

## NEED OF THE STUDY

### Implication of the outcome of the present study

Regards

Dr Devajit Nath

## REFERENCES

1. Organization, W. H. (2020) Clinical management of severe acute respiratory infection when novel coronavirus (2019-nCoV) infection is suspected: interim guidance, 28 January 2020, World Health Organization.

2. Lin Ling, Li Taisheng. Interpretation of the New Health Coronary Virus Infection Pneumonia Diagnosis and Treatment Program (Trial Version 5) [J]. Chinese Medical Journal, 2020, 100 (00): E001-E001.

3. Chan, J. F.-W., Yuan, S., Kok, K.-H., To, K. K.-W., Chu, H., Yang, J., Xing, F., Liu, J., Yip, C. C.-Y., and Poon, R. W.-S. (2020) A familial cluster of pneumonia associated with the 2019 novel coronavirus indicating person-to-person transmission: a study of a family cluster, The Lancet 395, 514–523.

4. Hoehl, S., Rabenau, H., Berger, A., Kortenbusch, M., Cinatl, J., Bojkova, D., Behrens, P., Böddinghaus, B., Götsch, U., and Naujoks, F. (2020) Evidence of SARS-CoV-2 infection in returning travelers from Wuhan, China, New England Journal of Medicine 382, 1278–1280.

5. Hu, Z., Song, C., Xu, C., Jin, G., Chen, Y., Xu, X., Ma, H., Chen, W., Lin, Y., and Zheng, Y. (2020) Clinical characteristics of 24 asymptomatic infections with COVID-19 screened among close contacts in Nanjing, China, Science China Life Sciences, 1–6.

6. Surveillances, V. (2020) The epidemiological characteristics of an outbreak of 2019 novel coronavirus diseases (COVID-19)—China, 2020, China CDC Weekly 2, 113–122.

7. Wang, Y., Liu, Y., Liu, L., Wang, X., Luo, N., and Ling, L. (2020) Clinical outcome of 55 asymptomatic cases at the time of hospital admission infected with SARS-Coronavirus-2 in Shenzhen, China, The Journal of infectious diseases.

8. Guan, W.-j., Ni, Z.-y., Hu, Y., Liang, W.-h., Ou, C.-q., He, J.-x., Liu, L., Shan, H., Lei, C.-l., and Hui, D. S. (2020) Clinical characteristics of coronavirus disease 2019 in China, New England Journal of Medicine.

9. Zhang, J.-j., Dong, X., Cao, Y.-y., Yuan, Y.-d., Yang, Y.-b., Yan, Y.-q., Akdis, C. A., and Gao, Y.-d. (2020) Clinical characteristics of 140 patients infected with SARS-CoV-2 in Wuhan, China, Allergy.

10. Chen, N., Zhou, M., Dong, X., Qu, J., Gong, F., Han, Y., Qiu, Y., Wang, J., Liu, Y., and Wei, Y. (2020) Epidemiological and clinical characteristics of 99 cases of 2019 novel coronavirus pneumonia in Wuhan, China: a descriptive study, The Lancet 395, 507–513.

11. Badawi, A., and Ryoo, S. G. (2016) Prevalence of comorbidities in the Middle East respiratory syndrome coronavirus (MERS-CoV): a systematic review and meta-analysis, International Journal of Infectious Diseases 49, 129–133.

12. Channappanavar, R., Fett, C., Mack, M., Ten Eyck, P. P., Meyerholz, D. K., and Perlman, S. (2017) Sex-based differences in susceptibility to severe acute respiratory syndrome coronavirus infection, The Journal of Immunology 198, 4046–4053.

13. Jaillon, S., Berthenet, K., and Garlanda, C. (2017) Sexual dimorphism in innate immunity, Clinical reviews in allergy & immunology, 1–14.

14. Li, Q., Guan, X., Wu, P., Wang, X., Zhou, L., Tong, Y., Ren, R., Leung, K. S., Lau, E. H., and Wong, J. Y. (2020) Early transmission dynamics in Wuhan, China, of novel coronavirus–infected pneumonia, New England Journal of Medicine.

15. Lippi, G., and Plebani, M. (2020) Procalcitonin in patients with severe coronavirus disease 2019 (COVID-19): A meta-analysis, Clinica Chimica Acta; International Journal of Clinical Chemistry.

16. Huang, C., Wang, Y., Li, X., Ren, L., Zhao, J., Hu, Y., Zhang, L., Fan, G., Xu, J., and Gu, X. (2020) Clinical features of patients infected with 2019 novel coronavirus in Wuhan, China, The Lancet 395, 497–506.

17. Yang, X., Yu, Y., Xu, J., Shu, H., Liu, H., Wu, Y., Zhang, L., Yu, Z., Fang, M., and Yu, T. (2020) Clinical course and outcomes of critically ill patients with SARS-CoV-2 pneumonia in Wuhan, China: a single-centered, retrospective, observational study, The Lancet Respiratory Medicine.

18. Bai, Y., Yao, L., Wei, T., Tian, F., Jin, D.-Y., Chen, L., and Wang, M. (2020) Presumed asymptomatic carrier transmission of COVID-19, Jama.

19. Qin, C., Zhou, L., Hu, Z., Zhang, S., Yang, S., Tao, Y., Xie, C., Ma, K., Shang, K., and Wang, W. (2020) Dysregulation of immune response in patients with COVID-19 in Wuhan, China, Clinical Infectious Diseases.

20. Xu, X.-W., Wu, X.-X., Jiang, X.-G., Xu, K.-J., Ying, L.-J., Ma, C.-L., Li, S.-B., Wang, H.-Y., Zhang, S., and Gao, H.-N. (2020) Clinical findings in a group of patients infected with the 2019 novel coronavirus (SARS-Cov-2) outside of Wuhan, China: retrospective case series, bmj 368.

21. Li, Q., Ding, X., Xia, G., Geng, Z., Chen, F., Wang, L., and Wang, Z. (2020) A simple laboratory parameter facilitates early identification of COVID-19 patients, medRxiv.

22. Qu, R., Ling, Y., Zhang, Y. h., Wei, L. y., Chen, X., Li, X., Liu, X. y., Liu, H. m., Guo, Z., and Ren, H. (2020) Platelet-to-lymphocyte ratio is associated with prognosis in patients with Corona Virus Disease-19, Journal of Medical Virology.

23. Forget, P., Khalifa, C., Defour, J.-P., Latinne, D., Van Pel, M.-C., and De Kock, M. (2017) What is the normal value of the neutrophil-to-lymphocyte ratio?, BMC research notes 10, 12.

24. Liu, J., Liu, Y., Xiang, P., Pu, L., Xiong, H., Li, C., Zhang, M., Tan, J., Xu, Y., and Song, R. (2020) Neutrophil-to-lymphocyte ratio predicts severe illness patients with 2019 novel coronavirus in the early stage, MedRxiv.

25. Fan, B. E., Chong, V. C. L., Chan, S. S. W., Lim, G. H., Lim, K. G. E., Tan, G. B., Mucheli, S. S., Kuperan, P., and Ong, K. H. (2020) Hematologic parameters in patients with COVID-19 infection, American journal of hematology.

26. Chng, W. J., Lai, H., Earnest, A., and Kuperan, P. (2005) Haematological parameters in severe acute respiratory syndrome, Clinical & Laboratory Haematology 27, 15–20.

27. Lee, N., Hui, D., Wu, A., Chan, P., Cameron, P., Joynt, G. M., Ahuja, A., Yung, M. Y., Leung, C., and To, K. (2003) A major outbreak of severe acute respiratory syndrome in Hong Kong, New England Journal of Medicine 348, 1986–1994.

28. Wenzhong, L., and Hualan, L. COVID-19: Attacks the 1-Beta Chain of Hemoglobin and Captures the Porphyrin to Inhibit Human Heme Metabolism. ChemRxiv. 2020, Preprint. https://doi.org/10.26434/chemrxiv 11938173, v6.

